# COVID-19 and Acute Kidney Injury Requiring Kidney Replacement Therapy: A Bad Prognostic Sign

**DOI:** 10.1101/2020.05.08.20096040

**Authors:** Rahul Shekhar, Shubhra Upadhyay, Silvi Shah, Devika Kapuria

## Abstract

The development of acute kidney injury in patients with COVID-19 is estimated to about 0.5% from earlier studies from China. The incidence of AKI in patients with COIVID-19 in the largest inpatient series in the United States is 22.2%<u>^3^</u>. Development of AKI requiring kidney replacement therapy in hospitalized patients is a bad prognostic sign. Out of Fifty patients admitted to our hospital with COVID-19 13/50(26%) developed AKI. All patients required hospitalization in intensive care unit care and 12/13 required initiation of kidney replacement therapy. The median age was 41 years (31-85 years) and 50% were men. Common comorbidities were obesity (83%), diabetes (42%), and hypertension (25%). 10/12 (83%) patients were hypoxemic and required oxygen therapy. 11/12 (92%) patients required invasive ventilation. Majority of patients had elevated neutrophils counts (81.8%) and low lymphocyte counts (81.8%). All patients had chest x-ray findings suggestive of pneumonia. 11/12(91.6%) developed septic shock requiring vasopressors. Review of UA showed all patient (9/9) had active urine sediments with blood but 7/9 of them have sterile pyuria. At the end of study period, 1 patient remained hospitalized. 10/11(90%) patients died and one patient was discharged home with resolution of AKI. Median length of stay was 13 days. The exact mechanism of AKI is not well understood in COVID-19 but can be due to acute tubular necrosis due to septic shock because of cytokine storm in severe COVID-19 or direct invasion by SARS-CoV-2 on podocytes and proximal renal tubular cells. Our findings suggest poor prognosis despite continuous kidney replacement therapies in patients who develop AKI with COVID-19.

## Introduction

The novel coronavirus, SARS-CoV-2 causing acute respiratory idleness termed COVID-19 by the World Health Organization (WHO) has affected millions of people worldwide. It was formally declared as a pandemic by WHO on March 11th 2020. Originating in the city of Wuhan, Hubei province of China, within months, this highly contagious virus has spread to most continents of the world. In the United States (US) alone, it has affected more than one million people and is responsible for 60,999 deaths so far (1).

Here we present our early experience in treating patients who were admitted to hospital for COVID-19, who developed Acute Kidney Injury (AKI) and required kidney replacement therapy. Observational studies from China show the incidence of AKI to be 0.5% (2) (3) overall. The incidence in the largest inpatient series in the US is 22.2% (4). In our cohort we observed that development of AKI requiring kidney replacement therapy in hospitalized patients is a bad prognostic sign and is associated with poor outcomes

## Methods

Retrospective chart review of the first fifty patients admitted to our hospital between Feb 1^st^ and April 24^th^ was done and patients who were diagnosed with COVID-19 via RT-PCR form nasal swab and developed AKI were included in this study. We characterized patients by demographics, comorbid conditions, severity of illness, laboratory test, chest film, CT scan, ultrasonography of kidney, Intensive Care Unit (ICU) use and treatment given. Detailed chart review was done by two independent physicians (RS, SU). In-hospital mortality estimates were calculated for patients with discharge dispositions. The University of New Mexico institutional review board approved the project with a waiver of informed consent

## Results

Of these fifty patients 26% (13/50) had AKI and 92% (12/13) developed oliguria, requiring renal replacement therapy. All patients were diagnosed with severe COVID-19 disease requiring transfer to ICU at some point during hospitalization. The median age was 41 years (31-85; 50% men) (See supplementary table 1). Common comorbidities were obesity (83%), diabetes (42%) and hypertension (25%). 83% (10/12) patients were hypoxemic on arrival and required oxygen therapy but 92% (11/12) patients ended up requiring intubation and mechanical ventilation due to acute respiratory distress syndrome (ARDS). Majority of patients have elevated neutrophils counts (82%), low lymphocyte counts (81.8%) (See supplementary table 2). All patients had chest x-ray findings suggestive of pneumonia, and majority developed septic shock requiring vasopressors (11/12-92%). All patients (9/9) had active urine sediments with blood, protein and majority had sterile pyuria (7/9). None of the patients had contrast exposure prior to development of AKI, only 1 patient had a history of CKD and 1 patient had history of intermittent NSAIDS use before admission. At the end of study period 1 patient remained hospitalized, 90% (10/11) patients had died and 1 was discharged home with resolution of AKI without requiring kidney replacement therapy. Median length of stay was 13 days. Other complications include multiorgan failure with liver injury in 4/11(36%), thromboembolic events in 2/11(18%), and cardiac arrest in 2/11(18%) patients.

## Discussion

About one fourth of patients admitted to hospital for treatment of COVID-19 developed AKI and the majority of them required kidney replacement therapy. Majority of these patients required intubation and mechanical ventilation secondary to ARDS, developed septic shock requiring vasopressors and the majority of them died. The exact mechanism of AKI is not well understood in covid-19 but can be due to ATN due to septic shock because of cytokine storm in severe COVID-19 or direct invasion by SARS-CoV-2 on podocytes and proximal renal tubular cells (5). Our findings suggest poor prognosis despite kidney replacement therapy in patients who develop AKI with COVID-19 and clinicians should closely watch for development for AKI especially patients with severe disease.

## Data Availability

Data is collect by two independent MD RS, SU and stored in redcap in deidentified format

## Abbreviation

WHO: World Health Organization
US: United States
AKI: Acute Kidney Injury
Ct: Computed tomography
ICU: Intensive care unit
ARDS: Acute respiratory distress syndrome
CRRT: Continuous renal replacement therapy
HD: Hemodialysis

---

**Table 1:**
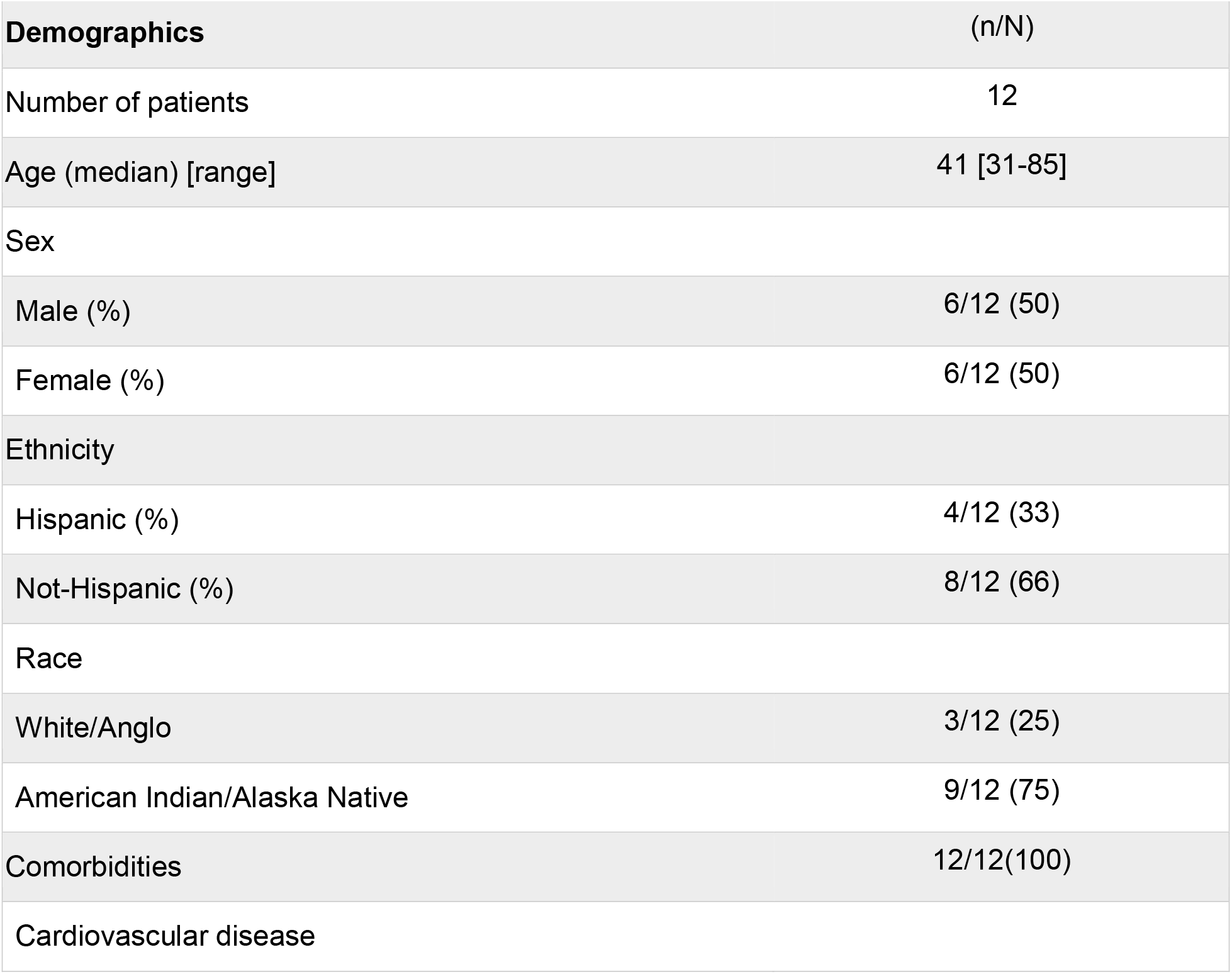
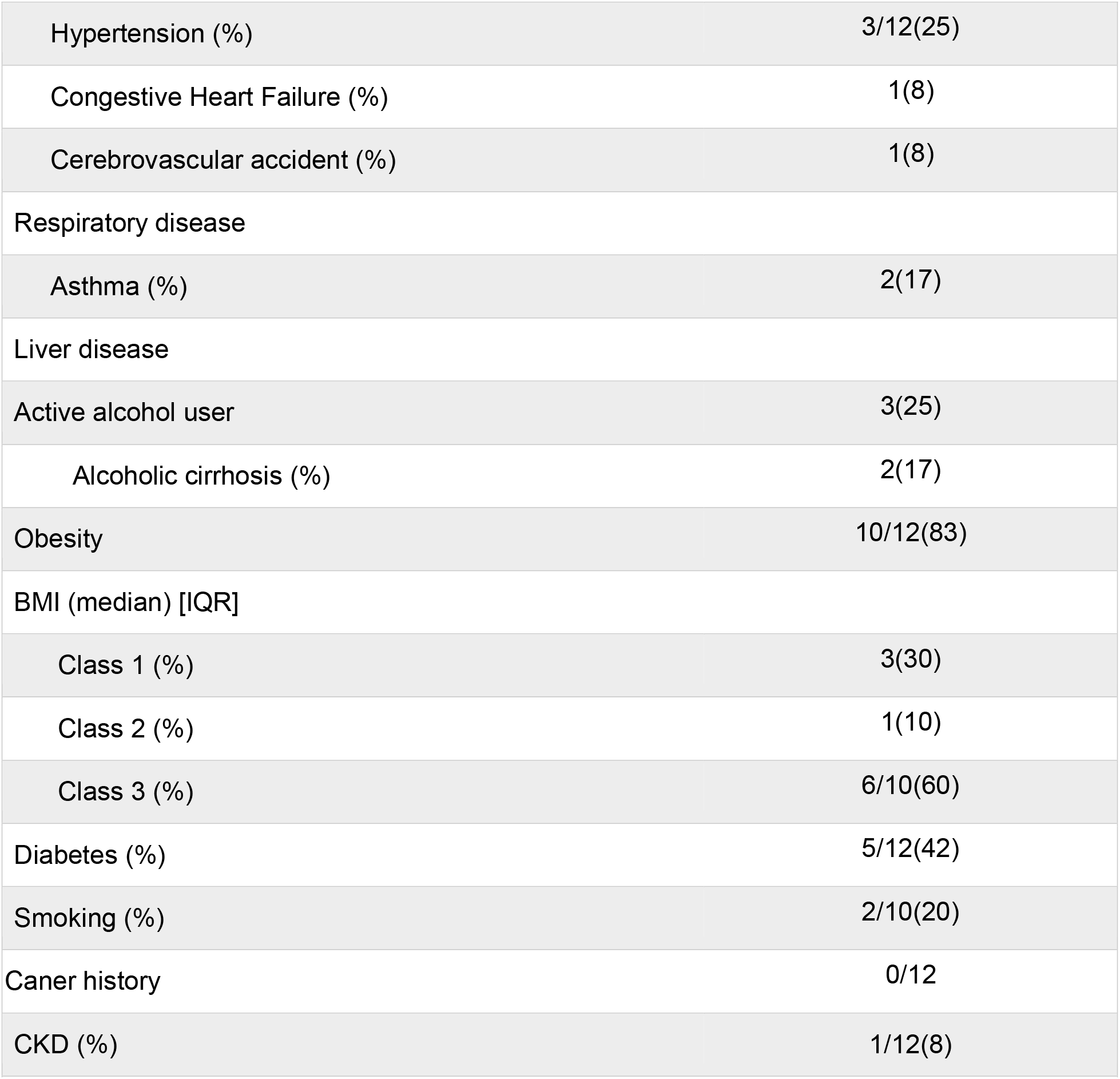
Baseline Characteristics

| **Table 1: Baseline Characteristics** |  |
| --- | --- |
| **Demographics** | **(n/N)** |
| Number of patients | 12 |
| Age (median) [range] | 41 [31-85] |
| Sex |  |
| Male (%) | 6/12 (50) |
| Female (%) | 6/12 (50) |
| Ethnicity |  |
| Hispanic (%) | 4/12 (33) |
| Not-Hispanic (%) | 8/12 (66) |
| Race |  |
| White/Anglo | 3/12 (25) |
| American Indian/Alaska Native | 9/12 (75) |
| Comorbidities | 12/12(100) |
| Cardiovascular disease |  |
| Hypertension (%) | 3/12(25) |
| Congestive Heart Failure (%) | 1(8) |
| Cerebrovascular accident (%) | 1(8) |
| Respiratory disease |  |
| Asthma (%) | 2(17) |
| Liver disease |  |
| Active alcohol user | 3(25) |
| Alcoholic cirrhosis (%) | 2(17) |
| Obesity | 10/12(83) |
| BMI (median) [IQR] |  |
| Class 1 (%) | 3(30) |
| Class 2 (%) | 1(10) |
| Class 3 (%) | 6/10(60) |
| Diabetes (%) | 5/12(42) |
| Smoking (%) | 2/10(20) |
| Cancer history | 0/12 |
| CKD (%) | 1/12(8) |

**Table 2:** Admission symptoms, vitals and labs

| All Patients |  |
| --- | --- |
| Admission vitals | (n/N) |
| Systolic BP (Median) [Mean] | 116(97-142) |
| Diastolic BP (median) [Range] | 74([43-84) |
| Mean Arterial pressure | 89(64-102) |
| O2 sats |  |
| <88% (%) | 10/12(83) |
| Febrile on admission (%) | 2/12(17) |
| Median Temperature * C | 36.65(35.2-38.3) |
| Initial oxygen treatment on arrival |  |
| On Room Air (%) | 1/12(8.3) |
| Nasal Cannula (%) | 4/12(25) |
| High flow / Nonrebreather (%) | 1/12 (8.3) |
| BIPAP (%) | 0/12(0) |
| Intubated (%) | 7/12(58.3) |
| Heart rate > 90 | 9/12(75) |
| Respiratory rate >20 (%) | 8/12(66.6) |
| Symptoms |  |
| Subjective fevers (%) | 6/7(85.7) |
| Cough (%) | 7/8(87.5) |
| Shortness of Breath (%) | 8/12(80) |
| Chest pain (%) | 1/2(50) |
| Nausea (%) | 2/4(50) |
| Diarrhea (%) | 4/6(66.7) |
| Vomiting (%) | 1/4(25) |
| Sore throat (%) | 2/2(100) |
| Abdominal pain (%) | 1/2(50) |
| constitutional symptoms (%) | 7/7(100) |
| Runny Nose (%) | 1/1(100) |
| **Laboratory Values (median) [IQR]** |  |
| WBC (median) [IQR] [4.0-11.0 E3/u] | (10.5) [6.60-33.30] |
| <4(%) | 0/12(0) |
| > 4 & < 11 (%) | 7/12(58.3%) |
| >11(%) | 5/12(41.7) |
| Neutrophils (median) [IQR] [ Normal Range1.8-7 10E3/uL] | (8.60) [5.50-23] |
| > 1.8 & < 7 (%) | 2/11(18.2) |
| >7 (%) | 9/11(81.8) |
| Lymphocytes (median) [IQR] [Normal range 1-3.4 10E3/uL] | (0.7) [.4 - 1] |
| < 1 (%) | 9/11(81.8) |
| >1 < 3.4 (%) | 2/11(18.2) |
| RBC [12.0-16.0 g/dl] | (11.7) [9.1-16.3] |
| Platelets [ Normal range 150 - 400 E3/UL] | (231) [54-377] |
| INR [ Normal Range 0.8-1.3 ratio] | (1.14) [.99 - 1.64] |
| D-Dimer [ Normal Range 0-500 ng/ml FEU] | (3075) [421-7650] |
| Ferritin [ Normal Range 18-340 ng/ml] | (1045) [220-1897] |
| CRP [ Normal Range <0.3 MG/DL] | (11.90) [.7 - 38.7] |
| LDH [ Normal Range 117-224 unit/l] | (614) [ 377-14152] |
| Lactate [ Normal Range 0.4-2.0 mmol/l] | (1.70) [1.2-13.3] |
| Procalcitonin [Normal Range <0.10 ng/ml] | (1.20) [.12-2.73] |
| A1C [ Normal Range 4.4-5.6 ] | (7.30) [4.0-15.60] |
| Na [ Normal Range 134 -144 mmol/L] | (134) [116-153] |
| K [ Normal Range3.5- 5.1 mmol/L] | (3.90) [3.0 - 4.90] |
| Chloride [ Normal Range 98-11 mmol/L] | (101) [80-121] |
| Bicarb | (20) [8-27] |
| BUN | (28.50) [8-225] |
| Creatinine [ Normal Range 0.50 - 1.40 mg/dl] | (3.07) [0.77-23.0] |
| Magnesium | (2.40) [1.70-3.40] |
| Phosphorus | (5.30)[2.50-16.70] |
| Abnormal Chest x ray | 12/12 (100) |
| MRSA Nasal swab (%) | 0/12(0) |
| Negative Blood culture | 11/12(91.6) |
| Hypotension/vasopressors (%) | 11/12(91.6) |
| pressures required / Median duration of pressure | 11/12(91.6)/ 8 days |
| Contrast exposure (%) | 0/12(0) |
| Nephrotoxic Drugs NSAIDS (%) | 1/12(8.3) |
| Hydronephrosis on Imaging (%) | 0/3(0) |
| Modality of treatment CRRT/HD/Conservative | CRRT 11/ 1 HD |
| Admission icu (%) | 12/12(100) |
| Mechanical ventilation (%) | 11/12(91.6) |
| UA positive for protein and blood (%) | 9/9(100) |
| UA positive wbc (%) | 7/9(77.7) |
| Median LOS in days (11/12) | 13 days (1-20days) |
| Outcome Death/Discharge/still admitted | 10 death/ 1 d/c home / 1 still admitted |
| Recovery (%) | 1/11(9) |

## Notes

### Competing Interest Statement

The authors have declared no competing interest.

### Funding Statement

Non Funded study

